# Motor-tasks fMRI BOLD activations in chronic stroke with residual hemiparesis in the upper extremity: a pre-neurofeedback baseline characterization

**DOI:** 10.64898/2026.04.15.26350962

**Authors:** Gabriele Varisco, Jeanette Plantin, Rita Almeida, Susanne Palmcrantz, Elaine Astrand

## Abstract

Stroke is the third leading cause of death and disability combined worldwide and often results in hemiparesis. Functional magnetic resonance imaging (fMRI) is a non-invasive technique used to investigate changes in brain activations during tasks aimed at restoring the lost motor function. Participants with chronic stroke and residual hemiparesis in the upper extremity were recruited for a clinical intervention that included neurofeedback training and fMRI sessions with motor-execution and motor-imagery tasks. The present study provides a baseline characterization of brain activations prior to neurofeedback training. Since lesion site and volume varied across participants, two fMRI preprocessing pipelines were applied. The first one was used for twelve participants with lesions restricted to a single hemisphere and for one participant with small secondary lesions in the contralesional hemisphere, whereas the second one was used for two participants with large bilateral lesions. These were followed by quality control measures and statistical analysis. First-level (i.e., single-participant) analysis returned the strongest and most extensive activation across participants during motor-execution tasks, with clusters identified in the ipsilesional parietal lobe, bilateral occipital lobes, and cerebellum after Family-Wise Error correction. Second-level (i.e., group-level) analysis involving participants who underwent the first fMRI preprocessing pipeline revealed a significant cluster in the cerebellum after False Discovery Rate correction. These results are consistent with previous studies involving participants with chronic stroke performing motor-tasks. Cerebellar recruitment observed consistently across participants could reflect compensatory mechanisms supporting motor control after stroke.

## 1 Introduction

According to the World Health Organization (WHO), stroke occurs when the blood flow reaching the brain is interrupted, resulting in the death of brain cells and serious complications [1]. Clinically, stroke can be classified as ischaemic, caused by occlusion of cerebral vessels, or hemorrhagic, caused by intracranial bleeding. Recent studies identify stroke as the third leading cause of death globally [1, 2, 3], as well as a major source of disability among adults [1, 4], resulting in significant socioeconomic costs [5]. Although not all clinical experts and associations agree with the definition proposed by the WHO [2], and a worldwide consensus has therefore not yet been reached, there is broad agreement on the clinical importance of stroke given its global burden.

Complications resulting from stroke can be separated into late medical (e.g., post-stroke seizures, urinary incontinence), musculoskeletal (e.g., hemiparesis, often presenting with weakness, spasticity, and impaired voluntary movements), and emotional (e.g., depression, anxiety, social isolation) [6]. Consequences include heightened neuroplasticity, often consisting of reorganization of brain functions through a lateralization of brain activity towards the intact hemisphere and/or functional reorganization within the lesioned hemisphere, shown for instance related to motor function [7].

Different non-invasive techniques have been adopted in past studies to investigate the brain activity in the presence of stroke, including electroencephalography (EEG) [8, 9], and functional magnetic resonance imaging (fMRI) [10]. fMRI, in particular, is used to analyze local changes in the blood-oxygen-level-dependent (BOLD) signal [11], which indirectly reflects neural activity and differs across tasks and experimental conditions. Additionally, different therapeutic strategies have been implemented aiming to recover the lost functions [12], many of which are tailored to the specific deficits and involve intensive, repetitive practice. An emerging strategy revolves around the use of neurofeedback [9, 13]. By employing sensors to record cortical activity while participants with stroke engage in specific tasks, it is possible to provide online feedback reflecting functionally relevant brain activity. This type of neurofeedback training is aimed at promoting neural plasticity through repeated practice and reinforcement of desired brain patterns.

Within our clinical intervention, we recruited a cohort of participants with chronic stroke (i.e., at least six months after the onset of stroke) with residual hemiparesis in the upper extremity. They participated in a series of EEG, fMRI, clinical assessments, and neurofeedback training sessions, with the overall objective of supporting the recovery of their motor function. Within this context, the present study aims to: (1) describe the fMRI preprocessing pipelines adopted to address challenges posed by stroke-related lesions; and (2) derive a baseline characterization of motor-task activations from the fMRI sessions conducted prior to neurofeedback training.

## 2 Methods

### 2.1 Dataset

This study was conducted in accordance with the Declaration of Helsinki. Approval to recruit participants and perform the clinical intervention was granted by both the Swedish Ethical Review Authority (Dnr 2022-00713-01) and the Swedish Medical Products Agency (CIV-21-10-037847). The study was registered as a clinical trial at clinicaltrials.gov (NCT04847089). All the included participants provided written informed consent prior to inclusion.

Following approval, a total of sixteen participants with chronic stroke and residual hemiparesis in the upper extremity were recruited by clinicians at Danderyd Hospital in Stockholm, Sweden. Specifically, eleven participants had an ischaemic stroke and five had a hemorrhagic stroke, all affecting brain regions supplied by the middle cerebral artery. One participant presented small secondary lesions in the contralesional hemisphere, caused by a non-stroke-related pathology, that did not overlap with voxels identified by mirroring the primary lesions, caused by stroke, across the mid-sagittal plane. In contrast, two participants presented large bilateral lesions, including secondary lesions in the contralesional hemisphere.

Stroke severity, assessed with the National Institutes of Health Stroke Scale (NIHSS), ranged from mild to moderate, with a mean score of 4. Hand motor impairment, assessed using the Fugl-Meyer Assessment for the Upper Extremity (FMA-UE), indicated moderate to severe impairment, with a mean score of 24. Additional information regarding the participants is provided in Table 1.

**Table 1:** Characteristics for the sixteen included participants with chronic stroke, reported as mean (min–max) for continuous variables and absolute numbers with percentages for categorical variables. Abbreviations: National Institutes of Health Stroke Scale (NIHSS), Fugl-Meyer Assessment for the Upper Extremity (FMA-UE).

| Characteristic | Values |
| --- | --- |
| Age (years at the time of inclusion) | 51 (28-73) |
| Sex | 11 (69%) male, 5 (31%) female |
| Stroke origin | 11 (69%) ischaemic, 5 (31%) haemorrhagic |
| Time from stroke onset to inclusion (months) | 33 (12-146) |
| Stroke severity (NIHSS) | 4 (0-10) |
| Hand motor impairment (FMA-UE) | 24 (11-46) |

### 2.2 Clinical intervention

The clinical intervention period spanned 10 weeks, during which each participant participated in a series of sessions, which are illustrated in Fig. 1 and reflecting those described in a previous study performed by our group [9]. A detailed description of sessions other than those using fMRI is beyond the scope of this study.

**Figure 1:**
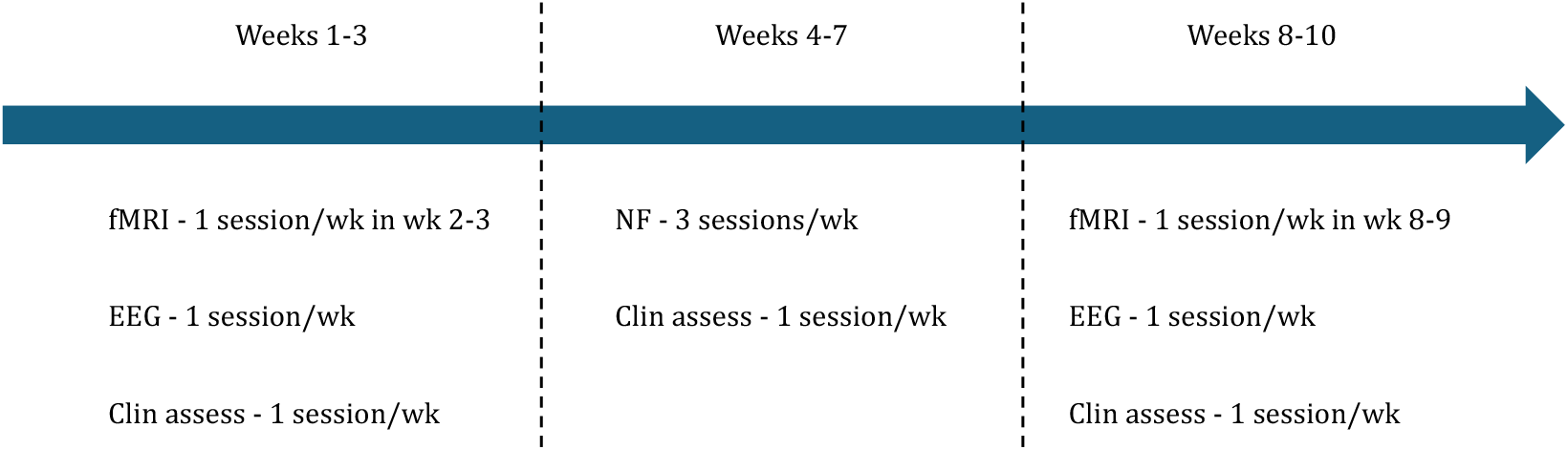
Clinical intervention period and scheduled functional magnetic resonance imaging (fMRI), electroencephalography (EEG), clinical assessments (Clin assess), and neurofeedback (NF) training sessions for the included participants with chronic stroke.

Four fMRI sessions were planned for each participant, respectively during week 2, 3, 8, and 9 of the clinical intervention period. Such planning was selected to enable the assessment of changes in participants’ condition before and after the neurofeedback training sessions. All fMRI sessions were recorded at Stockholm University Brain Imaging Centre in Stockholm, Sweden, using a Siemens MAGNETOM Prisma 3T MRI scanner, with a 20 channel head coil. During each fMRI session, three runs were acquired, consisting of two motor-task runs and one resting-state run.

All fMRI volumes were collected using gradient echo EPI, with voxel size of 2 × 2 × 2 mm, repetition time (TR) of 2090 ms, FOV = 200 mm, TE = 30 ms, in-plane acceleration GRAPPA 2, and multiband factor 2. During each motor-task run, participants first received instructions and then were presented with visual and auditory stimuli requesting them to perform one of five motor-tasks at each time: motor-execution of (1) grasping or (2) releasing an object (i.e., MEG and MER), motorimagery of (3) grasping or (4) releasing an object (i.e., MIG and MIR), and (5) idling/rest. With regards to motor-imagery tasks, in particular, participants were asked to mentally simulate the sensation of opening or closing their hand once and then maintain the imagined position [9]. During each motor-task run, MEG, MER, MIG, and MIR were performed four times each, whereas idling/rest was performed eight times. Within each motor-task run, the first presentation of a motor-imagery task always followed the presentation of the corresponding motor-execution task (i.e., MEG was followed by MIG, MER was followed by MIR). All subsequent motor-tasks were presented in a randomized order to maintain unpredictability for the participants, with idling/rest trials randomly interspersed among the other motor-tasks. Each motor-task had a similar duration, between 12 and 14 s.

Additionally, anatomical T1-weighted volumes were collected with MPrage TR = 2300 ms, TE = 2.98 ms, FOV = 256 mm, and voxel size = 1 × 1 × 1 mm, and T2-weighted volumes with FLAIR TR = 5000 ms, TE = 388 ms, FOV = 256 mm, and voxel size = 1 × 1 × 1 mm. T1-weighted volumes obtained during the first session for each participant were subsequently reviewed by a clinical expert from our authors’ group (J. P.), who manually derived lesion masks in MRIcron (Neuroimaging Tools and Resources Collaboratory, 2019) by annotating lesion voxels with a value of one.

### 2.3 Preprocessing

fMRI preprocessing, defined as a set of standardized transformations applied to reduce artifacts, correct acquisition-related effects, and spatially align volumes across participants to enable second-level (i.e., group-level) analysis, was then performed using SPM (v25; Wellcome Centre for Human Neuroimaging, University College London, United Kingdom), a MATLAB (R2024b, The MathWorks, Natick, Massachusetts, United States) toolbox [14].

However, the standard fMRI preprocessing pipeline performed with this toolbox does not account for the presence of lesions within fMRI and anatomical MRI volumes, which can introduce distortions that compromise the subsequent analyses. The Clinical Toolbox working within SPM addresses this issue by applying enantiomorphic lesion filling by mirroring voxels from the contralesional hemisphere [15]. This approach minimizes deformation during normalisation in lesioned brains, but its use remains limited to cases without large bilateral or symmetrical lesions. In such instances, the lesion-filled anatomical MRI volume provide sufficiently complete and symmetrical tissue maps to allow subsequent processing with the Diffeomorphic Anatomical Registration Through Exponentiated Lie algebra (DARTEL) algorithm [16]. This iteratively aligns tissue maps to build a group template, computes nonlinear deformation fields, and applies them to normalise fMRI volumes to both the group template and standard space. In the presence of large bilateral or symmetrical lesions, a practical alternative provided by SPM is the normalisation algorithm described in [17], often referred to as the Old Normalisation algorithm, which may however yield less anatomically precise normalisation compared to other more recent algorithms. Two separated fMRI preprocessing pipelines, shown in Fig. 2, were therefore used in this study.

**Figure 2:**
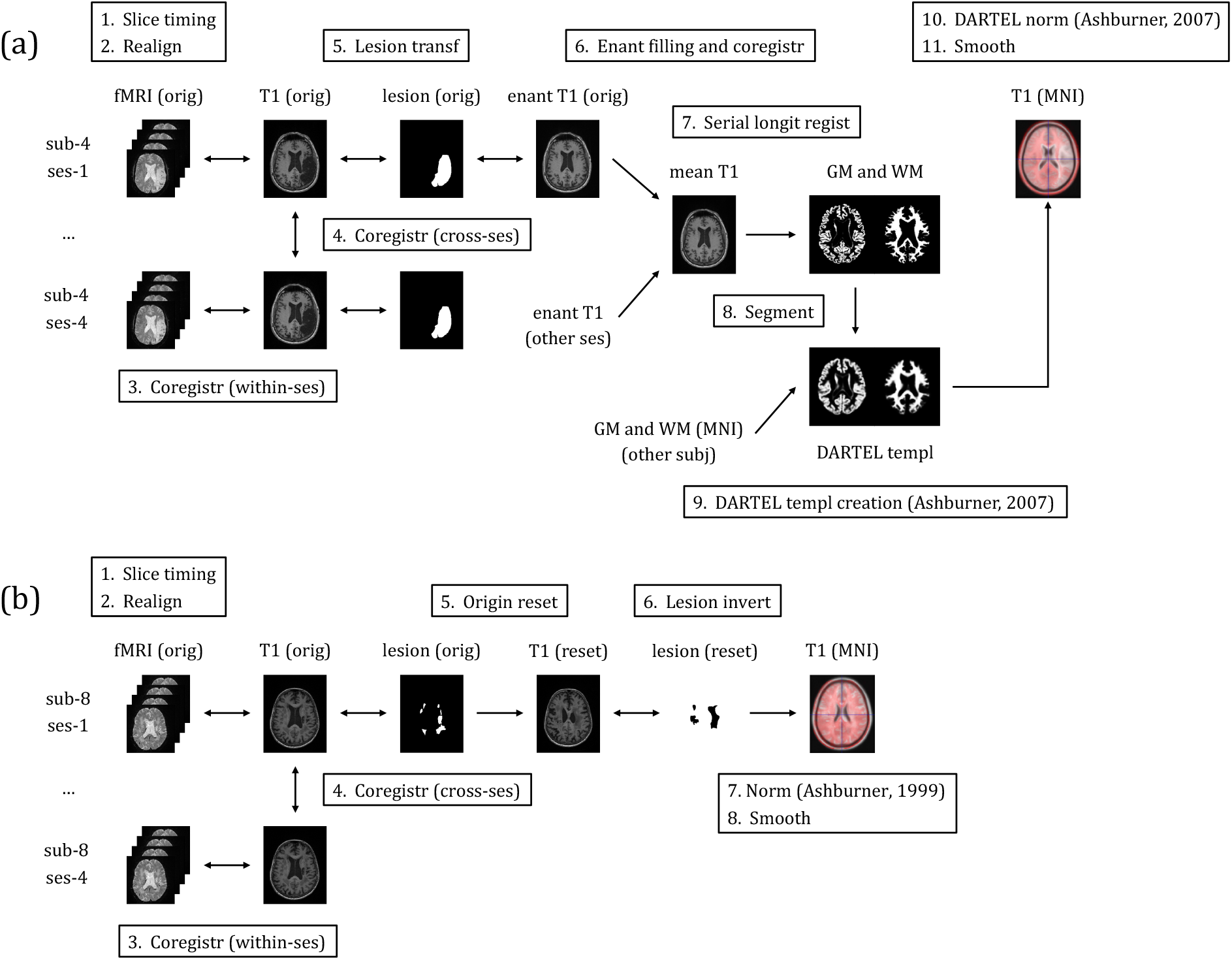
fMRI preprocessing pipelines used for participants: (a) with lesions restricted to a single hemisphere or with small secondary lesions in the contralesional hemisphere, and (b) with large bilateral lesions.

The fMRI preprocessing pipeline for participants with lesions restricted to a single hemisphere or with small secondary lesions in the contralesional hemisphere included: (1) slice timing correction; (2) realignment; (3) within-session coregistration of the fMRI volumes with the anatomical MRI volume from the same session; (4) cross-session coregistration, to align fMRI volumes and anatomical MRI volumes from all sessions to the anatomical MRI volume from the first session; (5) lesion mask transfer from the first session to following sessions; (6) enantiomorphic lesion filling of anatomical MRI volumes and cross-session coregistration, to provide optimal alignment with the anatomical MRI volume from the first session; (7) Serial Longitudinal Registration, to compute a mean anatomical MRI volume per participant; (8) segmentation, to map the mean anatomical MRI volume to a standard space and separate gray matter and white matter tissue maps; (9) DARTEL template creation [16], considering tissue maps from all participants and computation of nonlinear deformation fields; (10) DARTEL normalisation [16], to map fMRI volumes, anatomical MRI volumes, and lesion masks to standard space; and (11) spatial smoothing of the fMRI volumes with an isotropic Gaussian kernel with 6 mm full width at half maximum.

The fMRI preprocessing pipeline for participants with large bilateral lesions included the same initial four steps described for the previous pipeline followed by: (5) manual origin reset at the anterior commissure, to ensure correct alignment for subsequent spatial normalisation; (6) lesion mask inversion, to convert lesion voxels value to zero and non-lesioned voxels value to one; (7) use of the normalisation algorithm described in [17], together with the inverted lesion mask, to map fMRI volumes, anatomical MRI volumes, and lesion masks to standard space; and (8) spatial smoothing with an isotropic Gaussian kernel with 6 mm full width at half maximum.

Following both fMRI preprocessing pipelines and in line with previous studies [10], fMRI volumes characterized by right-sided primary lesions, caused by stroke, were flipped along the mid-sagittal plane so that all primary lesions appeared on the left side, allowing group-level contrasts to be interpreted consistently as ipsilesional or contralesional to the lesioned hemisphere.

fMRI preprocessing outputs were manually inspected at each step to ensure correct alignment and confirm that no substantial deformation was introduced. Alignment with the T1-weighted template provided by SPM was used to verify the accuracy of normalisation. Additionally, since differences between fMRI preprocessing pipelines may lead to small but systematic variations in voxel-wise alignment in standard space due to the the use of distinct spatial normalisation approaches, which differ in deformation model, template construction, and registration accuracy [16, 18], the normalised fMRI and anatomical volumes obtained from the two fMRI preprocessing pipelines were directly compared prior to performing second-level analysis.

### 2.4 Quality control

Quality control was carried out using a two-step approach. First, MRIcroGL [19] was used to perform a visual inspection of fMRI volumes that had not undergone fMRI preprocessing to look for artifacts, such as interference patterns caused by hardware-related instabilities and ghosting resulting from gradient imperfections or phase-encoding errors. This task was jointly performed by two authors (G.V. and J.P.), allowing immediate discussion regarding the identified artifacts.

Second, the Artifact Detection Tools (ART) (Neuroimaging Tools and Resources Collaboratory, 2015) [20] working within SPM was used to identify outliers, defined as fMRI volumes derived from realignment and affected by significant head motion during their acquisition. More conservative thresholds than others considered for other clinical populations (i.e., 9 standard deviations for the z-normalised global signal and 2 mm for the translational and rotational motion parameters derived from realignment [21]) were applied. Specifically, thresholds were set to 5 standard deviations for the z-normalised global brain signal and 1.5 mm for the translational and rotational motion parameters. This choice allowed to balance the number of fMRI volumes to exclude from first-level (i.e., single-participant) analysis while avoiding overfitting.

### 2.5 First- and second-level analysis

In this study, all motor-task runs from the two initial fMRI sessions planned for each participant (i.e., up to four motor-task runs) were modeled together and analyzed to establish a baseline characterization of motor-task activations.

First-level analysis was performed in SPM by defining a general linear model (GLM) for each participant [14, 22]. This included regressors for each motor-task run for MEG, MER, MIG, and MIR. Each task was specified with onsets and durations in seconds and modeled by convolution with the canonical hemodynamic response function, including temporal and dispersion derivatives. Addi-tional regressors included for each motor-task run modeled the six realignment parameters, derived from realignment, the artifacts and outliers, derived from quality control, as well as an intercept term. Lesion masking allowed to exclude lesioned voxels from the estimation. Additionally, any motor-task run with more than 15% outliers was excluded, resulting therefore in some participants having less than the initial four motor-task runs included in the analysis.

After GLM estimation, different effects of interest were investigated using dedicated t-contrasts. These included the average effect of performing any of the motor-tasks (i.e., MEG, MER, MIG, and MIR) relative to baseline (*‘any task* > *baseline’*), as well as the average effects of performing motor-execution tasks alone (*‘motor execution* > *baseline’*) and motor-imagery tasks alone relative to baseline (*‘motor imagery* > *baseline’*). The derived statistical maps for each participant were assessed using Family-Wise Error (FWE) correction for multiple comparisons with a threshold of p < 0.05.

Second-level analysis was then performed to assess significant activations at the group level (i.e., across participants) by means of a One-sample t-test [14]. Significance was assessed both at an uncorrected voxel-wise threshold of p < 0.001 as well as using False Discovery Rate (FDR) correction for multiple comparisons.

## 3 Results

One participant was excluded from the analysis due to non-attendance at scheduled sessions and insufficient data across the clinical intervention. Two additional participants withdrew from the clinical intervention but completed the first two fMRI sessions. Therefore, they were included in the present study to derive a baseline characterization of motor-task activations from the fMRI sessions conducted prior to neurofeedback training.

Twelve participants with lesions confined to a single hemisphere and the single participant with small secondary lesions in the contralesional hemisphere (largest bounding-box diameter ≈ 13 mm), underwent the fMRI preprocessing pipeline that included enantiomorphic lesion filling and DARTEL normalisation. Two participants presenting large bilateral lesions underwent the fMRI preprocessing pipeline that relied on the normalisation algorithm described in [17]. Normalisation to standard space using either fMRI preprocessing pipeline did not produce substantial deformation in any of included participants, although the resulting normalised fMRI and anatomical volumes showed small spatial discrepancies between the two fMRI preprocessing pipelines. For this reason, participants who underwent the former fMRI preprocessing pipeline constituted the full dataset for second-level analysis, whereas the two participants with bilateral lesions were considered only at the first-level and qualitatively compared with the activation patterns obtained from the main dataset.

Quality control inspection performed using MRIcroGL identified ghosting in some of the included runs. Nonetheless, these artefacts appeared outside of the brain volumes and did not contaminate brain voxel values. Therefore, no motor-task run was excluded from the consequent analysis due to ghosting. In contrast, the use of ART with the selected thresholds led to the exclusion of a total of 12 motor-task runs, including all four motor-task runs from one participant, who was subsequently excluded from further analysis.

First-level analysis revealed substantial inter-subject variability in the strength and extent of activations, reflecting different degrees of responsiveness across the cohort. The statistical map *‘any task* > *baseline’* returned significant activations in all but two participants. In these two participants only a very small number of isolated voxels reached significance when considering FWE correction with a threshold of p < 0.05, consistent with noise-level or non-robust activations. Because this result reflects the average response across all motor-execution and motor-imagery tasks, further insights into task-specific patterns were derived by examining the corresponding t-contrasts.

The statistical map *‘motor execution* > *baseline’* returned the strongest and most extensive activation patterns throughout the cohort, with only one participant showing no suprathreshold activation when considering FWE correction with a threshold of p < 0.05. The distribution of activations varied across individuals, but clusters were most commonly observed in the ipsilesional parietal lobe, the bilateral occipital lobes, and the cerebellum, reflecting the involvement of somatosensory, visual, and cerebellar motor-related processing regions. Additionally, all participants who showed activations in the occipital lobes also exhibited concurrent activation in the parietal lobe. The statistical map from a sample participant (i.e., sub-2), showing activations in all three identified brain regions, is shown in Fig. 3a.

**Figure 3:**
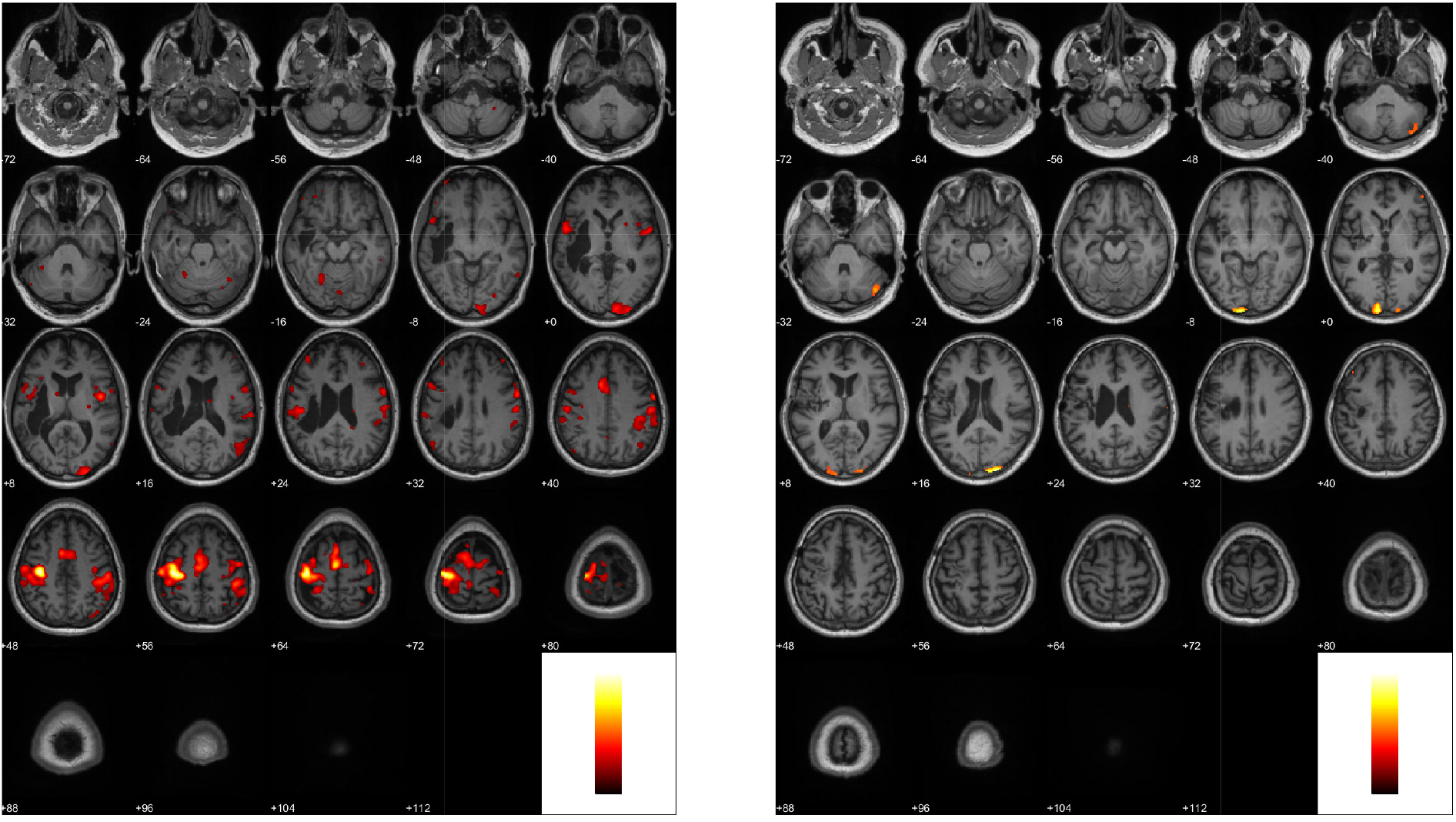
First-level analysis results: (a) statistical map *‘motor execution* > *baseline’* derived from sub-2, and (b) statistical map *‘motor imagery* > *baseline’* derived from sub-17. In both cases, significance was assessed using Family-Wise Error (FWE) correction with a threshold of p < 0.05.

In contrast, the statistical map *‘motor imagery* > *baseline’* returned the weakest overall responses, with three participants showing no suprathreshold activation except for a few isolated voxels when considering FWE correction with a threshold of p < 0.05. Among the remaining participants, activations tended to be more spatially restricted compared to what was noticed with the previous statistical map, typically involving the parietal lobe, with lateralization varying by participant, the bilateral occipital lobes, and the cerebellum. Notably, isolated activations in the occipital lobes without a concurrent parietal involvement were found in two participants, a pattern that was not observed with the previous statistical map. The statistical map from a sample participant (i.e., sub-17), showing this restricted activation pattern, is shown in Fig. 3b.

With regard to the effects of using different fMRI preprocessing pipelines, no atypical or distinguishable activation patterns were observed across any of the investigated statistical maps in the two participants who underwent the fMRI preprocessing pipeline that relied on the normalisation algorithm described in [17]. These statistical maps were found to be consistent with the overall variability seen in the rest of the included cohort.

Second-level analysis computed considering all participants who underwent the fMRI preprocessing pipeline that included enantiomorphic lesion filling and DARTEL normalisation revealed only a limited number of suprathreshold voxels across the investigated statistical maps when significance was assessed using an uncorrected voxel-wise threshold of p < 0.001. The only statistical map that showed a significant effect after applying FDR correction, as shown in Fig. 4a, was *‘motor execution* > *baseline’*, which revealed a group-level cluster in the cerebellum. The results found using the statistical map *‘motor imagery* > *baseline’* when using an uncorrected voxel-wise threshold of p < 0.001, reported in Fig. 4b, showed only isolated suprathreshold voxels, often located at the borders between the sulci and the gyri or along ventricular edges, a pattern that is consistent with the presence of noise or residual motion artifacts, especially since these voxels did not survive correction for multiple comparisons.

**Figure 4:**
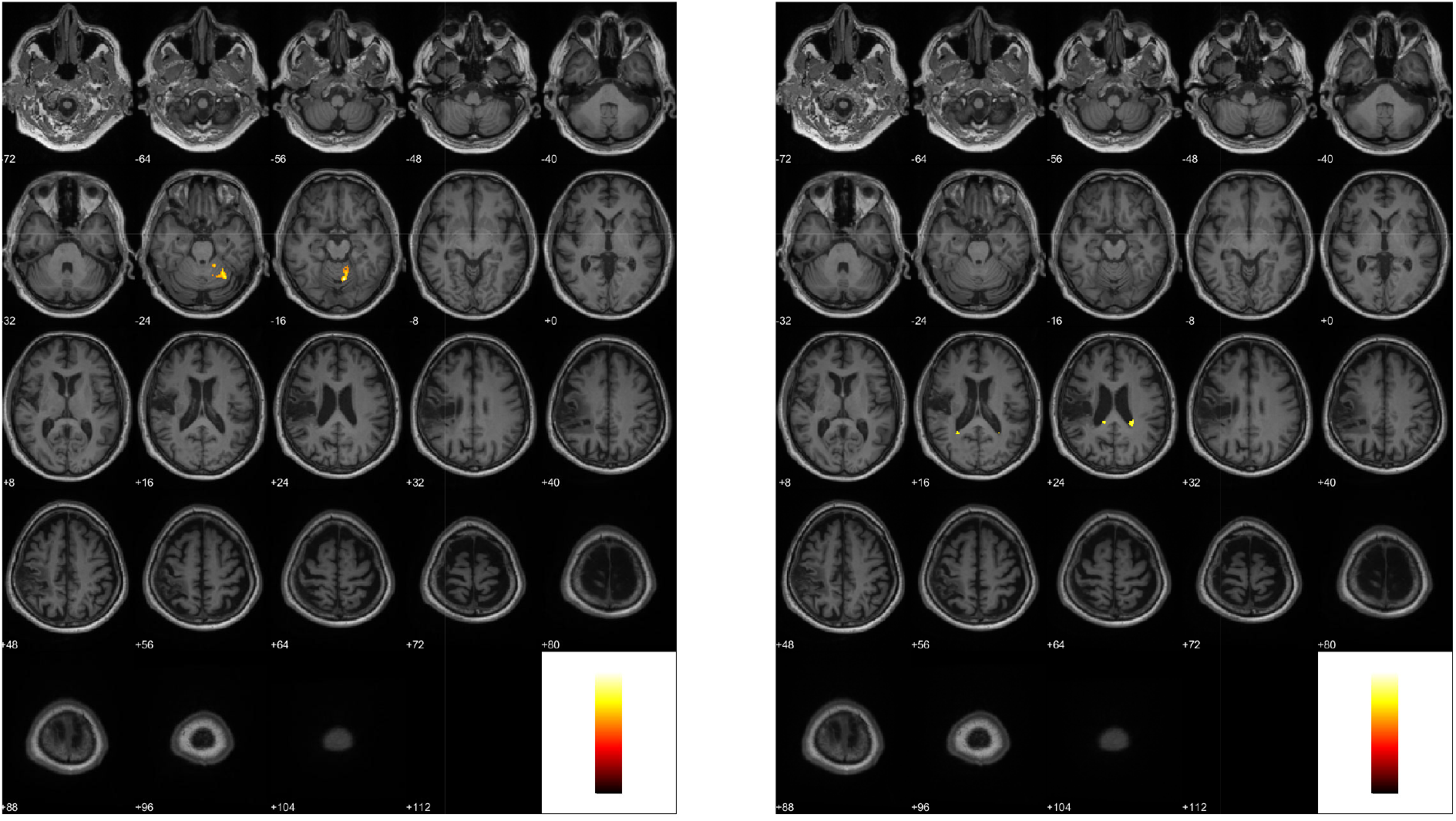
Second-level analysis results: (a) statistical map *‘motor execution* > *baseline’* derived from all participants that underwent the fMRI preprocessing pipeline that included enantiomorphic lesion filling and DARTEL normalisation, where significance was assessed using False Discovery Rate (FDR) correction, and (b) statistical map *‘motor imagery* > *baseline’* derived from the same participant group, where significance was assessed using an uncorrected voxel-wise threshold of p < 0.001. Both statistical maps are overlaid on an anatomical MRI volume derived from sub-4.

## 4 Discussion

Within our clinical intervention, we recruited a cohort of participants with chronic stroke and residual hemiparesis in the upper extremity who participated in neurofeedback training alongside other sessions to improve their motor function [9]. In the present study, we performed a baseline evaluation of motor-task activations derived from fMRI sessions, which will serve as a reference for assessing changes associated with neurofeedback training in future studies.

We applied two dedicated fMRI preprocessing pipelines, respectively to participants with lesions restricted to a single hemisphere or with small secondary lesions in the contralesional hemisphere, and with large bilateral lesions, in both cases considering the presence of multiple runs per participant and a single lesion mask. Both fMRI preprocessing pipelines relied on SPM, a widely used toolbox working in MATLAB that provides robust statistical modelling and advanced normalisation methods that are validated in clinical populations [14]. The fMRI preprocessing pipeline that included enantiomorphic lesion filling and DARTEL normalisation [16], applied to the former group of participants and representing the majority of our dataset, offered different advantages. First, it provided a fully automated workflow, without requiring a manual origin reset at the anterior commissure, thereby reducing user-dependent variability and ensuring reproducibility. Second, since no manual placement at the anterior commissure was required, it avoided the normalisation inaccuracies that can arise from imprecise selection of this anatomical landmark and the need for multiple iterations to achieve a satisfactory result, issues that made the fMRI preprocessing pipeline that relied on the normalisation algorithm described in [17] more time-consuming and less reliable. Third, since DARTEL relies on a nonlinear, high-dimensional registration approach [16], estimating far more deformation parameters than the normalisation algorithm described in [17], it resulted in improved inter-subject alignment. Nevertheless, only small spatial discrepancies were observed when considering fMRI and anatomical MRI volumes derived from the two fMRI preprocessing pipelines, which were not characterized by substantial deformations in either case. These differences, however, were sufficient to prevent including all participants into second-level analysis.

The consequent inclusion of quality control measures allowed to exclude any possible influence of artifacts from the included runs. Selecting appropriate thresholds for identifying outliers and applying them to the standard deviations for the z-normalised global signal and the translational and rotational motion parameters derived from realignment represent a crucial step especially when analyzing resting-state runs [23], where excessive motion can substantially bias connectivity estimates. In the present study, however, despite only task-based runs were investigated, it was decided to adopt quite conservative thresholds compared to other studies involving clinical populations [21] since it was observed that the absence of adequate motion filtering would have resulted in visibly blurred activation patterns (results not shown), thereby compromising the interpretability of the statistical maps derived from first- and second-level analysis. At the same time, thresholds were not selected to be as strict as those typically used in resting-state analysis, since motor-tasks inherently induce some degree of movement [24], and participants with stroke can exhibit additional involuntary movements, tremor, as well as reduced ability to maintain a stable posture [25].

Statistical maps derived from first-level analysis are consistent with those found in previous fMRI studies involving participants with chronic stroke. In particular, activations in the parietal and occipital lobes have been reported when performing motor-tasks [26] and visual-tasks [27], respectively. The presence of activations in the occipital lobes in our dataset is therefore expected, since our participants received visual instructions requesting them to perform motor-tasks. Stronger parietal activations due to motor-tasks were observed in the ipsilesional hemisphere, consistent with previous studies involving participants with chronic stroke [28]. In contrast, participants with acute stroke are more often characterized by predominant contralesional parietal activations, with longitudinal neuroimaging studies indicating a gradual shift toward ipsilesional recruitment over time, which is also associated with improved motor outcomes [29].

The present study identified activations in the cerebellum in statistical maps derived both from first- and second-level analysis, highlighting the involvement of a brain structure that plays a key role in movement coordination, sensorimotor integration, and motor adaptation. Cerebellar recruitment during motor-tasks in participants with stroke has been consistently reported, with a recent systematic review showing compensatory hyperactivation of the cerebellum relative to healthy controls [30], and could possibly reflect increased demands on motor coordination, error correction, and adaptive control mechanisms in response to impaired cortical motor networks. On the other hand, the statistical maps derived from second-level analysis did not reveal a significant global activation in the parietal and occipital lobes, likely due to inter-subject variability in the spatial distribution of activations and to the presence of large lesions affecting these regions across participants.

Another result found in this study is that motor-imagery tasks led to statistical maps with weaker activations compared to those found with motor-execution tasks. In two participants, in particular, activations were restricted to occipital lobes without parietal involvement, which may reflect a greater reliance on visual processing rather than effective kinesthetic imagery during motor-imagery tasks. Despite undergoing a different fMRI preprocessing pipeline, the two participants with large bilateral lesions did not show statistical maps characterized by atypical or qualitatively distinct motor-task activations compared to the rest of the cohort, suggesting that the alternative fMRI preprocessing pipeline did not introduce systematic differences.

This study presents different strengths as well as limitations. A first major strength relates to the structure of the clinical intervention, which included multiple fMRI sessions and multiple motor-task runs per session both before and after the introduction of neurofeedback training sessions. This choice increased the amount of data available per participant and improved the reliability of first-level estimates, allowing the GLM to more robustly capture effects derived from motor-tasks by reducing noise and increasing statistical power.

A second strength lies in the comprehensive fMRI preprocessing strategy, which used two dedicated fMRI preprocessing pipelines, one for participants with lesions confined to a single hemisphere or with small secondary lesions in the contralesional hemisphere, and another for those with large bilateral lesions, combined with the inclusion of considered quality-control measures. This approach was essential for identifying outliers caused by head motion, which are common in clinical populations such as participants with chronic stroke and could otherwise lead to spatial blurring of activations and reduced interpretability of the results. Additionally, the investigation of different effects of interest through dedicated t-contrasts allowed a clear separation of neural responses associated with different motor-tasks.

A first limitation relates to the relatively small number of participants with chronic stroke who were included in our clinical intervention. This was primarily due to the stringent inclusion criteria required to identify suitable candidates (e.g., sufficient proficiency in the local language, and absence of additional neurological or medical conditions that could confound treatment effects), as well as the substantial time and resources required to complete the full clinical intervention, which spanned several weeks and involved repeated clinical assessments, fMRI acquisitions, and dedicated clinical personnel. Consequently, the further exclusion from second-level analysis of participants presenting large bilateral lesions further reduced the effective sample size. Future studies could address this issue by exploring alternative software solutions, such as fMRIPrep [31], to evaluate their capa-bilities for performing fMRI preprocessing in the presence of lesions of varying site and volume. A second limitation relates to the absence of a dedicated analysis of resting-state runs acquired prior to the introduction of neurofeedback training. Although these were collected as part of our clinical intervention, their inclusion was beyond the scope of the present study and will be addressed in future studies to complement the findings derived from task-based runs and provide a more comprehensive characterization of functional network changes found in our cohort of participants with chronic stroke.

## Data Availability

Due to strict ethical considerations, restrictions on data sharing apply.

## Acknowledgment

We would like to thank Prof. Jörgen Borg, MD, for his support in recruiting participants for our clinical intervention and for his constructive input during its planning and design. We also extend our thanks to Jonatan Tidare, Martin Johansson-Alvarez, and Jenny Sylvén for their assistance with the EEG and fMRI data collection.

